# Patch2Self denoising of diffusion MRI in the cervical spinal cord improves intra-cord contrast, signal modelling, repeatability, and feature conspicuity

**DOI:** 10.1101/2021.10.04.21264389

**Authors:** Kurt G. Schilling, Shreyas Fadnavis, Joshua Batson, Mereze Visagie, Anna J.E. Combes, Colin D. McKnight, Francesca Bagnato, Eleftherios Garyfallidis, Bennett A. Landman, Seth A. Smith, Kristin P. O’Grady

## Abstract

Quantitative diffusion MRI (dMRI) is a promising technique for evaluating the spinal cord in health and disease. However, low signal-to-noise ratio (SNR) can impede interpretation and quantification of these images. The purpose of this study is to evaluate a denoising approach, Patch2Self, to improve the quality, reliability, and accuracy of quantitative diffusion MRI of the spinal cord. Patch2Self is a self-supervised learning-based denoising method that leverages statistical independence of noise to suppress signal components strictly originating from random fluctuations. We conduct three experiments to validate the denoising performance of Patch2Self on clinical-quality, single-shell dMRI acquisitions with a small number of gradient directions: 1) inter-session scan-rescan in healthy volunteers to evaluate enhancements in image contrast and model fitting; 2) repeated intra-session scans in a healthy volunteer to compare signal averaging to Patch2Self; and 3) assessment of spinal cord lesion conspicuity in a multiple sclerosis group. We find that Patch2Self improves intra-cord contrast, signal modeling, SNR, and lesion conspicuity within the spinal cord. This denoising approach holds promise for facilitating reliable diffusion measurements in the spinal cord to investigate biological and pathological processes.

## Introduction

Quantitative diffusion MRI (dMRI) is a promising tool to study the tissue microstructure of the spinal cord in health and disease. To date, the most commonly utilized dMRI technique is diffusion tensor imaging (DTI). DTI provides quantitative indices including fractional anisotropy (FA), mean diffusivity (MD), axial diffusivity (AD), and radial diffusivity (RD), which have been shown to be sensitive to tissue properties such as axon density, axonal injury, and degree of myelination (Beaulieu, 2002). In addition to DTI, a growing number of more advanced microstructure models, or multi-compartment models, of diffusion have been applied to the spinal cord (Duval et al., 2015; Grussu et al., 2019; Grussu et al., 2017; Grussu et al., 2016; Moccia et al., 2019; Saliani et al., 2017; Schilling et al., 2019), improving pathological specificity to tissue damage. For example, in multiple sclerosis (MS), a condition featured by a complex interplay among inflammation, demyelination and axonal loss, DTI indices have been shown to correlate well with clinical measures of disability (Moccia et al., 2019). Further, advanced measures of neurite density, compartment diffusivities, and axonal disorganization have shown sensitivity in identifying abnormal changes in MS spinal cord lesions, as well as in normal-appearing white matter (WM) (By et al., 2017, 2018; Cohen-Adad, 2018; Grussu et al., 2016; Grussu et al., 2015; Schilling et al., 2019), and suggest an expanding use for diffusion-derived metrics in MS clinical practice and trials.

Yet, spinal cord dMRI on clinical scanners is challenging. The spinal cord is a thin, complex structure requiring relatively high spatial resolution for adequate anatomic depiction. Spinal cord imaging is also complicated by artifacts from motion and local susceptibility, resulting in lower SNR and precision in quantitative analysis. At the same time, in vivo imaging has scan time constraints, which limits the number of diffusion weighted images (DWIs) that can be acquired, and requires fundamental tradeoffs with image resolution and image quality. Thus, improving image quality is imperative to facilitate both research investigations and clinical application of spinal cord dMRI.

Towards this end, several denoising approaches have been applied to dMRI data, typically in the brain, in order to improve signal-to-noise ratio (SNR) and contrast-to-noise ratio (CNR), as well as reproducibility and precision of diffusion-derived features. Most denoising methods for dMRI typically utilize one of three approaches (Buades et al., 2005; Coupe et al., 2008; Elad and Aharon, 2006; Knoll et al., 2011; Manjon et al., 2013; Rudin et al., 1992): (1) they may assume that the signal is intrinsically low-rank (local PCA, MP-PCA) and can be denoised through dimensionality reduction, (2) they may assume that the signal patterns may be replicated throughout the tissue (non-local means) and can be denoised through averaging of similar signals, or (3) assume that the signal is locally smooth (total variation norm) and can be denoised through spatial smoothing. Recently, the Marchenko-Pastur PCA denoising (MP-PCA) has been proven to significantly enhance SNR and improve parameter maps in the brain and spinal cord (Grussu et al., 2020; Veraart et al., 2016a; Veraart et al., 2016b). However, this technique exploits data redundancy (i.e. multiple DWIs or multi-contrast datasets) which may not exist with time-limited datasets and low numbers of DWIs. A recently proposed image denoiser, Patch2Self, is unique in that it makes no assumptions on the structure of the signal, and only assumes that noise is random and uncorrelated across DWIs. Recent work (Fadnavis et al., 2020) has shown promising results in suppressing noise and preserving anatomical detail in the brain; however, the effectiveness of this method in the spinal cord has not been investigated.

Thus, the aim of this study is to evaluate the efficacy of Patch2Self denoising on clinical-quality dMRI data in the cervical spinal cord in healthy volunteers and in patients with MS. We first utilize a scan-rescan dataset to evaluate the effects of Patch2Self denoising on intra-cord contrast, fitting of diffusion models, and diffusion reproducibility. Next, we compare Patch2Self against a long-time many-average experiment. Finally, we apply the algorithm to data from an MS cohort, and assess lesion conspicuity before and after denoising. We hypothesize that Patch2Self denoising would lead to improved CNR, model-fitting, reproducibility, SNR, and lesion conspicuity in spinal cord diffusion images.

## Materials and Methods

### Datasets

This study contained 3 data cohorts, designated A, B, and C, that were used to evaluate different effects of the denoising process (**Figure 1**).

**Figure 1.**
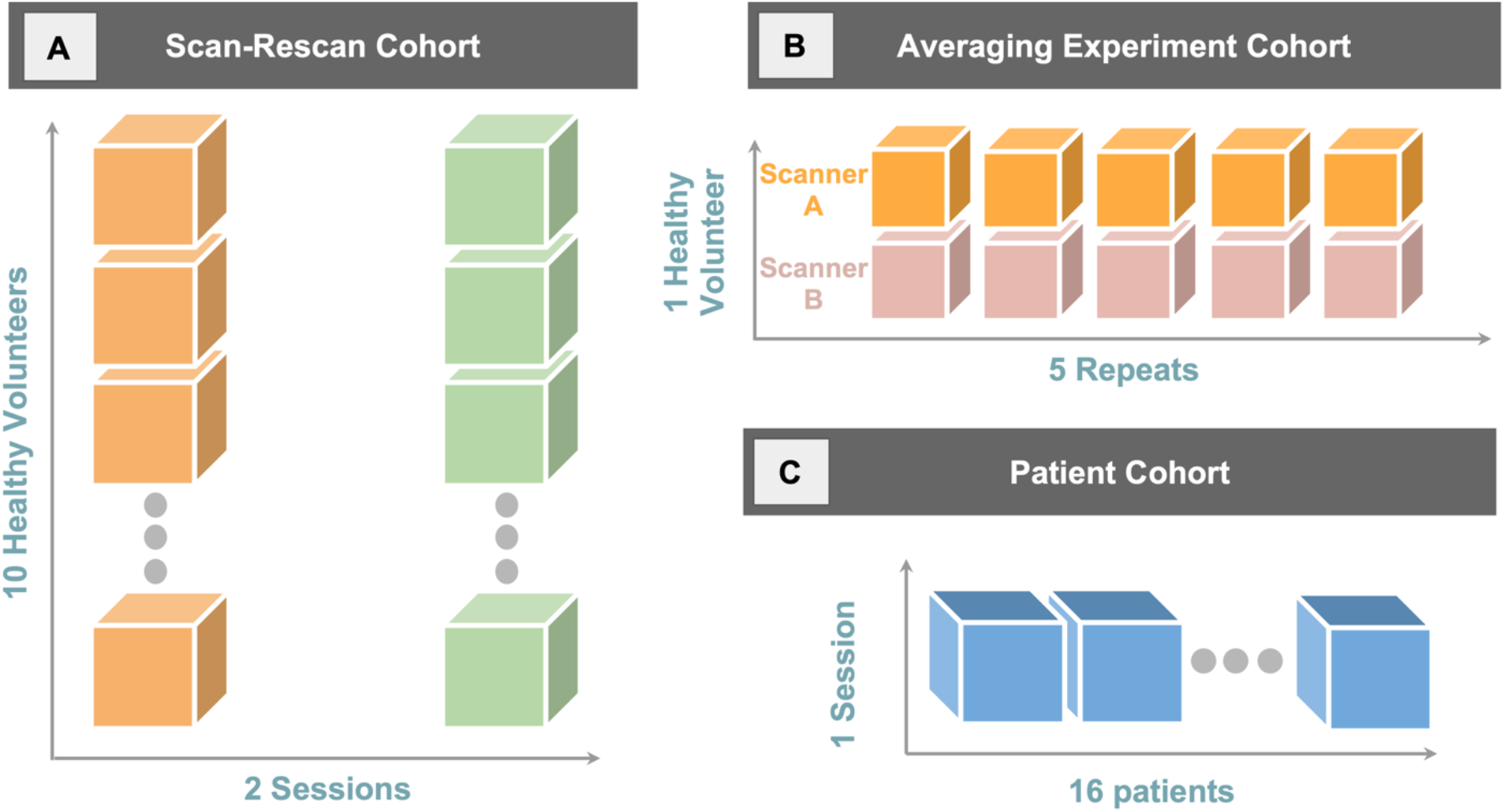
Three cohorts used to evaluate Patch2Self denoising in the spinal cord. (A) The scan-rescan cohort was composed of 10 healthy volunteers scanned twice, and was used to visualize denoising results, quantify CNR, and investigate inter-session reproducibility. (B) The averaging experiment cohort consisted of 1 healthy volunteer on two scanners, where the acquisition was repeated 5 times, which allowed comparison of denoising to averaging many datasets. (C) The patient cohort consistent of 16 pwRRMS, with one scan per patient, that allowed visualization and assessment of image quality and lesion conspicuity after denoising.

Cohort A was a scan-rescan dataset, consisting of N=10 healthy controls (HCs), aged between 22 and 40 years, 5 females (F) and 5 males (M), with two sessions acquired 3-5 months apart. This dataset was used to visualize effects of denoising, quantify CNR, and investigate inter-session repeatability of DTI-derived metrics. Imaging was performed using a 3T Philips Elition MR scanner with 2-channel transmit and a dStream neurovascular coil (Philips) for reception.

Cohort B included one female HC in their 30s with 5 repeated acquisitions within one session on the same 3T Philips Elition MR scanner as Cohort A. This experiment allowed us to compare a denoised dataset to one with improved anatomy visualization and increased SNR due to signal averaging. This acquisition was repeated on a different 3T Philips dStream Ingenia MR scanner at the same site in order to show robustness to potential scanner effects.

Cohort C consisted of N=16 people with relapsing-remitting MS (pwRRMS) (20-42 years old, 9F/7M, Expanded Disability Status Scale scores 0-1.5) with one session per patient. This cohort was used to assess the effects of denoising on image quality, contrast, and lesion conspicuity. Imaging was performed using the same 3T Philips Elition MR scanner as in Cohort A.

### Acquisition

All cohorts utilized the same imaging acquisition protocol. For each session, a high-resolution (0.65×0.65×5 mm^3^) multi-slice, multi-echo gradient echo (mFFE) anatomical image (Held et al., 2003) was acquired (TR/TE/ΔTE = 700/8.0/9.2 ms, α = 28 degrees, number of slices = 14, 6:12 minutes) for co-registration and to serve as a structural reference image. The diffusion sequence consisted of a cardiac-triggered (using a peripheral pulse oximeter with a delay set to 127 ms) spin echo sequence with single-shot echo planar imaging (EPI) readout with the following parameters: TR/TE = 5 beats (∼5000 ms)/77 ms, resolution = 1.1×1.1mm^2^, slice thickness = 5 mm, FOV = 80 × 57.5 × 70 mm, SENSE (RL) = 1.8, partial Fourier = 0.693, and NEX = 3, with 14 axial slices (time ∼6-8 minutes depending on heart rate). All images were centered between the C3 and C4 levels. Reduced field-of-view was applied using an outer volume suppression technique (Wilm et al., 2007) and fat suppression was achieved using SPIR. A single-shell acquisition was used with 15 diffusion-weighted directions at b = 750s/mm.

### Processing

The processing pipeline is shown in **Figure 2**. Diffusion data from each session were motion-corrected using the Spinal Cord Toolbox (SCT; (De Leener et al., 2017)), and cropped to minimize the field-of-view outside of the cord. Next, Patch2Self was applied to the cropped volumes. Quantitative DTI parameters of FA, AD, RD and MD were calculated using FSL’s linear least squares algorithm from both raw and denoised data. All model-fitting was performed in native space. The mean diffusion-weighted image for each scan was segmented to obtain a cord mask, and registered to the structural image (mFFE) using SCT, and transforms were saved in order to overlay/visualize data in the desired space. Finally, the structural image was registered to the PAM50 spinal cord template (De Leener et al., 2018) using SCT. The resulting inverse warp field was used to propagate template labels (WM, GM) to subjects’ native space for quantitative analysis.

**Figure 2.**
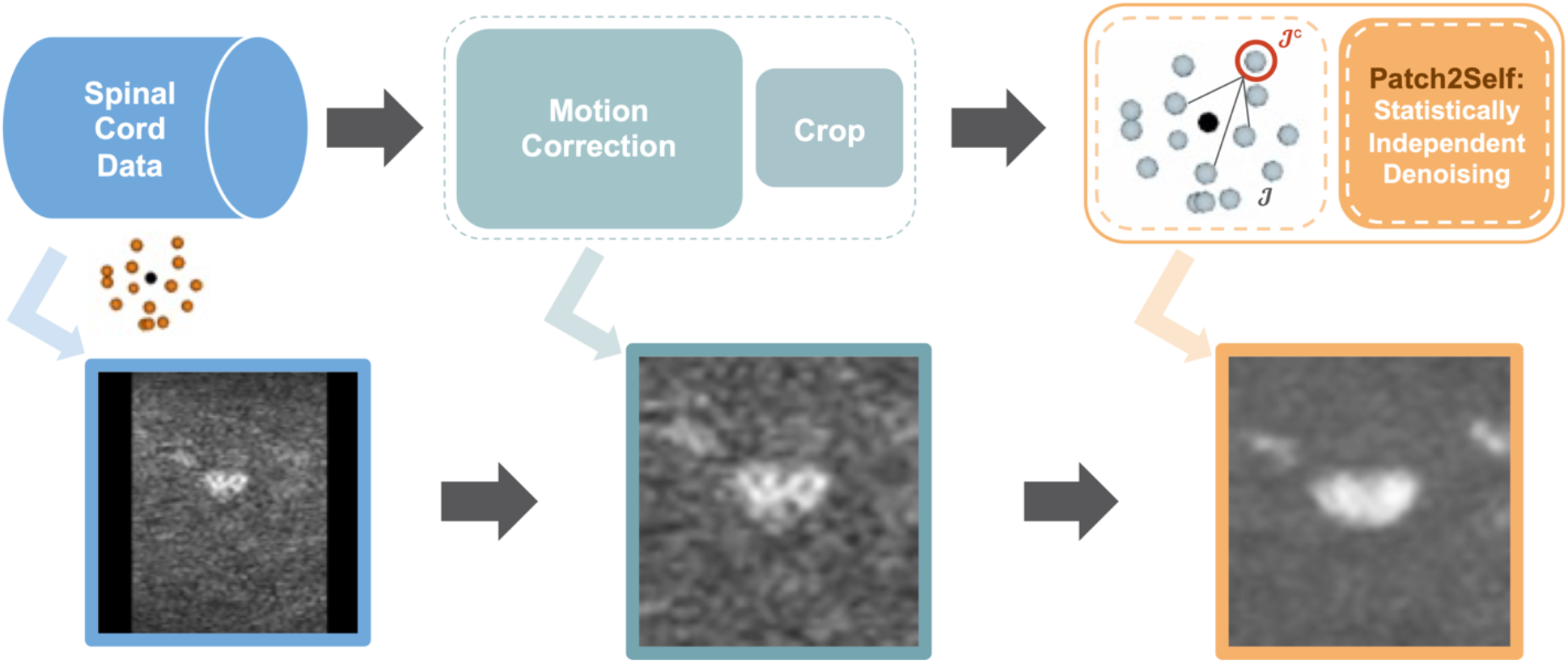
Image processing pipeline. Diffusion data were motion corrected using the Spinal Cord Toolbox, then Patch2Self (implemented in DIPY) was applied to cropped, motion-corrected volumes.

### Patch2Self Theory

The implementation of Patch2Self using DIPY 1.4.1 was employed to perform the denoising of all spinal cord data presented in this paper (Fadnavis et al., 2020). Anisotropic patches of radii [2, 2, 0] (units of mm) along the [x, y, z] directions of the image were used in order to deal with the anisotropic nature of the spinal cord image. Patch2Self can be deployed at any stage of the pre-processing pipeline since it does not rely on any assumptions on the signal, as long as the noise in each 3D volume of the data is statistically independent of the noise in the other 3D volumes. In order to deal with the motion inherent to spinal cord images, motion correction was done prior to performing the denoising.

Patch2Self proposes a novel way of using self-supervised learning to perform the denoising. Building on top of the J-invariance theory proposed in (Batson and Royer, 2019) and statistical independence in (Lehtinen et al., 2018), Patch2Self starts with extracting 3D patches from each 3D volume of the 4D diffusion MRI data. Thus, if the data had *n* volumes with *m* voxels and each patch has the radius *p*, the extracted patches would be: *(p* × *p* × *p)* × *m* × *n*. These extracted patches are then flattened to become 2D so that the resulting shape is: *mp*^*3*^ × *n* (denoted as 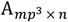). In the case of spinal cord, *p* is typically set to 1 or 2 depending on the dimensionality of the data. In this paper, *p* = 1 has been used for all the experiments. Next as per the theory suggested in Patch2Self, a J-invariant self-supervised loss is minimized between the held-out volume and the remaining *n* − 1 volumes. This training loss can be written as follows:

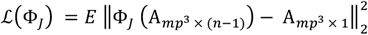

Here, Φ_*j*_ is the self-supervised loss that is minimized in the case of spinal cord data using the default linear regression model available in DIPY. Once the training is done, the same trained model is now used to predict the held out volume *J* using the trained function. The prediction obtained is the denoised volume and this procedure is applied iteratively on all 3D volumes of the 4D data. (Note: The held-out volume *J* is only used as the target for training and not in the design matrix 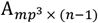.

This procedure can be seen as a q-space in-painting method (**Figure 2**, cartoon depictions) where each gradient direction is represented as a linear combination of the remaining gradient directions. Note that Patch2Self does not impose any signal properties such as smoothness, low-rank, signal repetition, etc. that are usually leveraged in commonly used denoising methods such as Total Variation, MP-PCA and NLMeans. Although it makes use of the redundancy of the data in q-space, Patch2Self does not make any explicit assumptions while learning the denoising function.

### Evaluation

#### Qualitative evaluation

All visualizations of DWIs and derived maps were shown in native diffusion space. The only exception to this was the Cohort C data, in which all data were transformed to subject anatomical space (mFFE) to facilitate lesion identification and comparison.

#### CNR calculation

To assess image contrast before and after denoising, we calculated the CNR between WM and gray matter (GM) in the raw DWIs. Contrast was calculated as the absolute value of the average signal intensity of the WM at C3 subtracted from the average signal intensity of the GM at C3. The noise standard deviation was calculated as the standard deviation of the signal of the CSF in the DWIs (this is because of the high diffusivity of CSF, which should result in noise in diffusion weighted images at this diffusion weighting). CNR was then calculated as contrast divided by noise standard deviation, quantified for all DWIs, and averaged. CNR was calculated for the first scan for all subjects in Cohort A and compared before and after denoising using a paired t-test.

#### Model fitting evaluation

In order to evaluate the goodness-of-fit of the DTI model to the data, we performed a per-voxel k-fold cross-validation. To do so, in each fold we held out a set of data points and predicted them after fitting the DTI model. This was done k times and a goodness-of-fit was computed relative to the S0 signal. For this paper, k=3 was used to perform the validation. The implementation used to compute the R^2^ is available in DIPY as per (Rokem et al., 2015) and (Hastie et al., 2009). Once an R^2^-score was obtained for each voxel, we summarized the improvement in the goodness-of-fit for the DTI model for all voxels in the data using box-plots.

#### Reproducibility

We assessed scan-rescan reproducibility of DTI-derived metrics, before and after denoising. To do this, for each subject, we calculated the mean value of four metrics (FA, MD, AD, and RD) in four regions-of-interest (WM of C3, WM of C4, GM of C3, and GM of C4). We then calculated the mean absolute error (MAE) as the absolute difference between scan and rescan. We visualize the average MAE for all regions and compare those before and after denoising using a paired t-test.

#### Comparison against multiple averages

Using Cohort B, we compared denoising using Patch2Self on a single dataset (acquisition time ∼7 minutes) to a dataset equivalent to multiple averages (up to ∼7×5 = 35 minutes). To do this, we calculated the voxel-wise SNR as the average DWI signal in a voxel divided by the noise standard deviation, calculated as the square root of the sum of squared residuals to a tensor fit. SNR was calculated for all voxels within the cord mask.

#### Lesion conspicuity evaluation

To quantitatively evaluate image quality, a trained neuroradiologist (C.M.) was presented with montages from Cohort C, of 3 randomly selected DWIs in 3 consecutive axial slices, along with the corresponding structural image. We note that this random selection of slices and DWIs does not guarantee a lesion in every image. In total, 160 images were presented in a random order (80 matching denoised and non-denoised), and the neuroradiologist was asked to provide a single grade for each montage “on a scale of 1-5 based on observed contrast, lesion conspicuity (if present), signal to noise, and presence/absence of artifacts”. This allowed a quantitative assessment of the effects of denoising on perceived image quality.

## Results

### Scan-Rescan Experiments

Cohort A was used to illustrate image quality after denoising on healthy controls, quantify CNR before and after denoising, assess improvements in signal modelling, and investigate scan-rescan reproducibility of quantitative DTI-derived measures.

Figure 3 shows qualitative results on two example subjects, comparing raw diffusion images and Patch2Self denoised images of the same slice. The first notable observation is that the noise variance is reduced, both within the structure and in background. Second, intra-cord contrast has qualitatively increased, with the GM “butterfly” shape more apparent, particularly in already low signal DWIs. Third, no notable artifacts nor spurious signals are visibly introduced.

**Figure 3:**
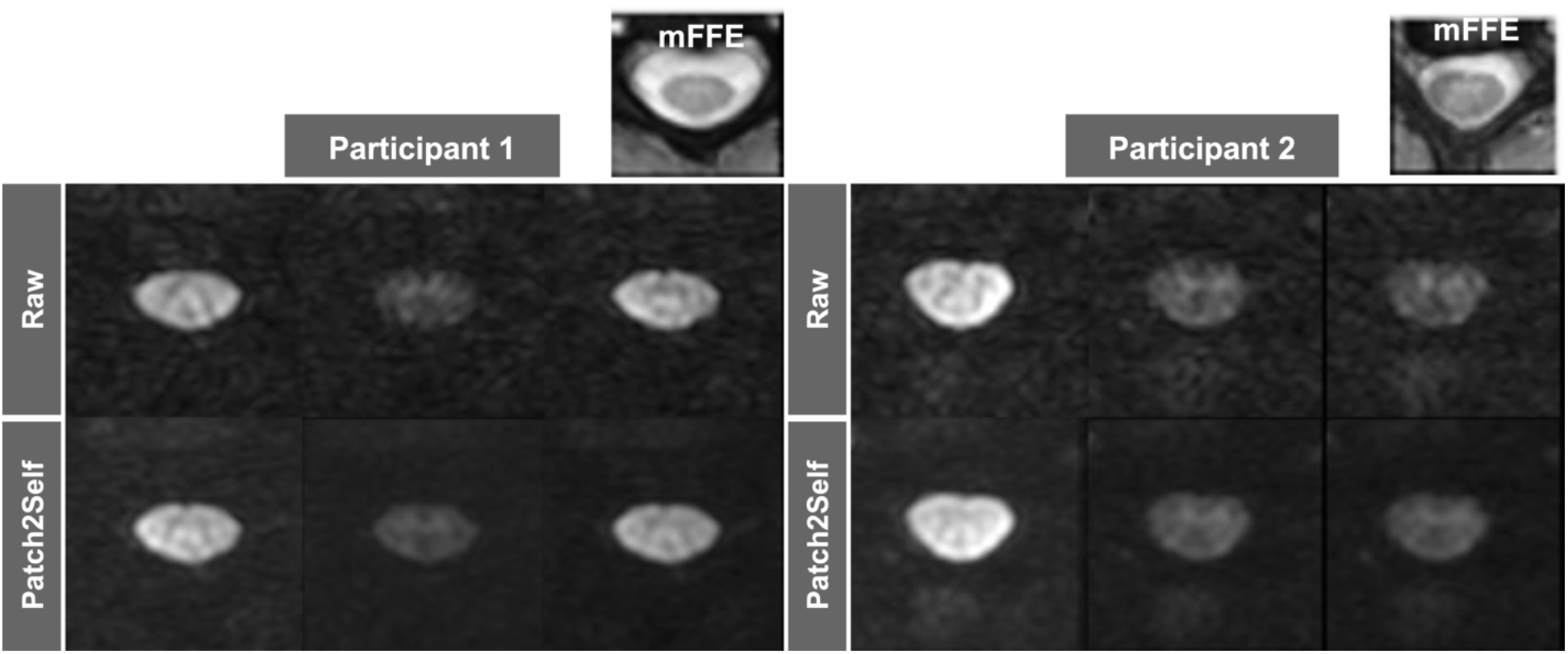
Denoising the diffusion-weighted images qualitatively improves tissue contrast, as shown for two healthy volunteers in Cohort A. Here, raw diffusion data is shown on top, and denoised on bottom.

Figure 4 quantifies the CNR between WM and GM regions of the cord, before (Raw) and after (Patch2Self) denoising, across all 10 subjects. In agreement with qualitative observations, the WM/GM CNR is significantly higher after denoising (p=1.1E-4; median CNR from 2.9 to 4.0) with an average 35% increase in CNR. Additionally, we found that the CNR of derived metrics (FA, MD, AD, RD) did not significantly increase after denoising (not shown), which highlights the absence of effect on the magnitude of derived indices in WM and GM.

**Figure 4:**
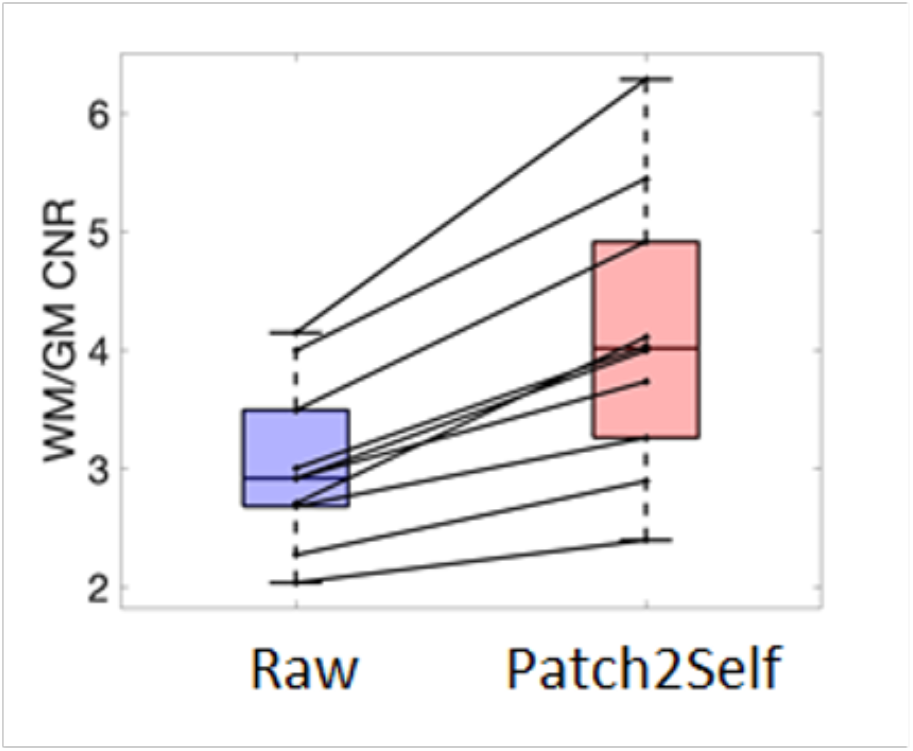
Patch2Self denoising increases the white matter to gray matter contrast to noise ratio (CNR) for 10 healthy volunteers in Cohort A. Increase in CNR is statistically significant (p<.001).

Figure 5 shows the improvement in the goodness-of-fit using the R^2^-scores computed per voxel using a cross-validation approach described above. In nearly all cases, R^2^ increases significantly, indicating a better fit to the data with the DTI model after denoising.

**Figure 5:**
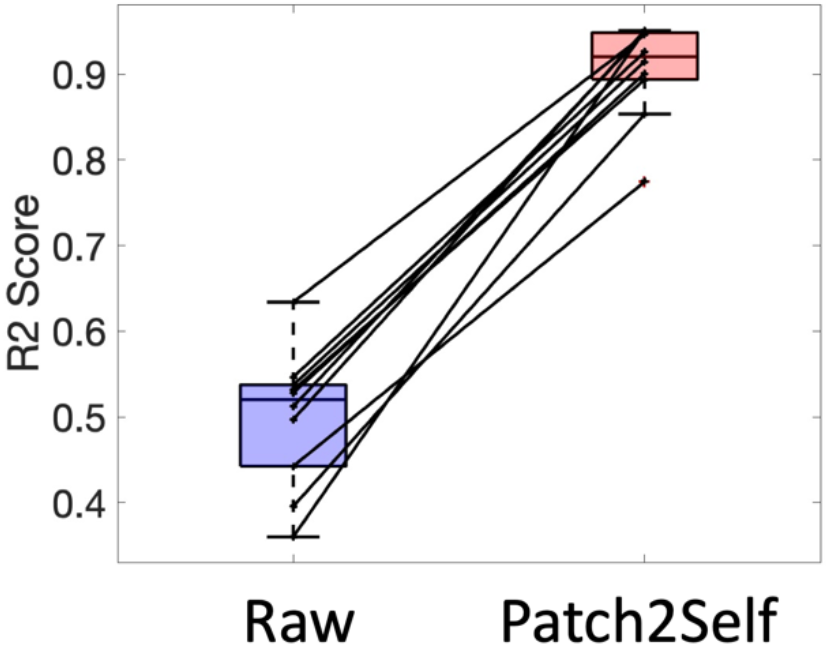
Goodness of fit of the DTI signal model (R^2^) increases significantly with denoising. Shown are mean R^2^ of the cord for 10 healthy volunteers in Cohort A. Increase in R^2^ is statistically significant (p<.001).

Finally, we assess scan-rescan reproducibility of DTI-derived metrics. **Figure 6** shows the MAE of FA, MD, AD, and RD before and after denoising, where a lower MAE indicates more reproducible measures across sessions. The MAE of FA (p=0.005) and AD (p=0.021) decreased (i.e., reproducibility increased) after denoising, whereas MD and RD showed no significant changes in reproducibility.

**Figure 6:**
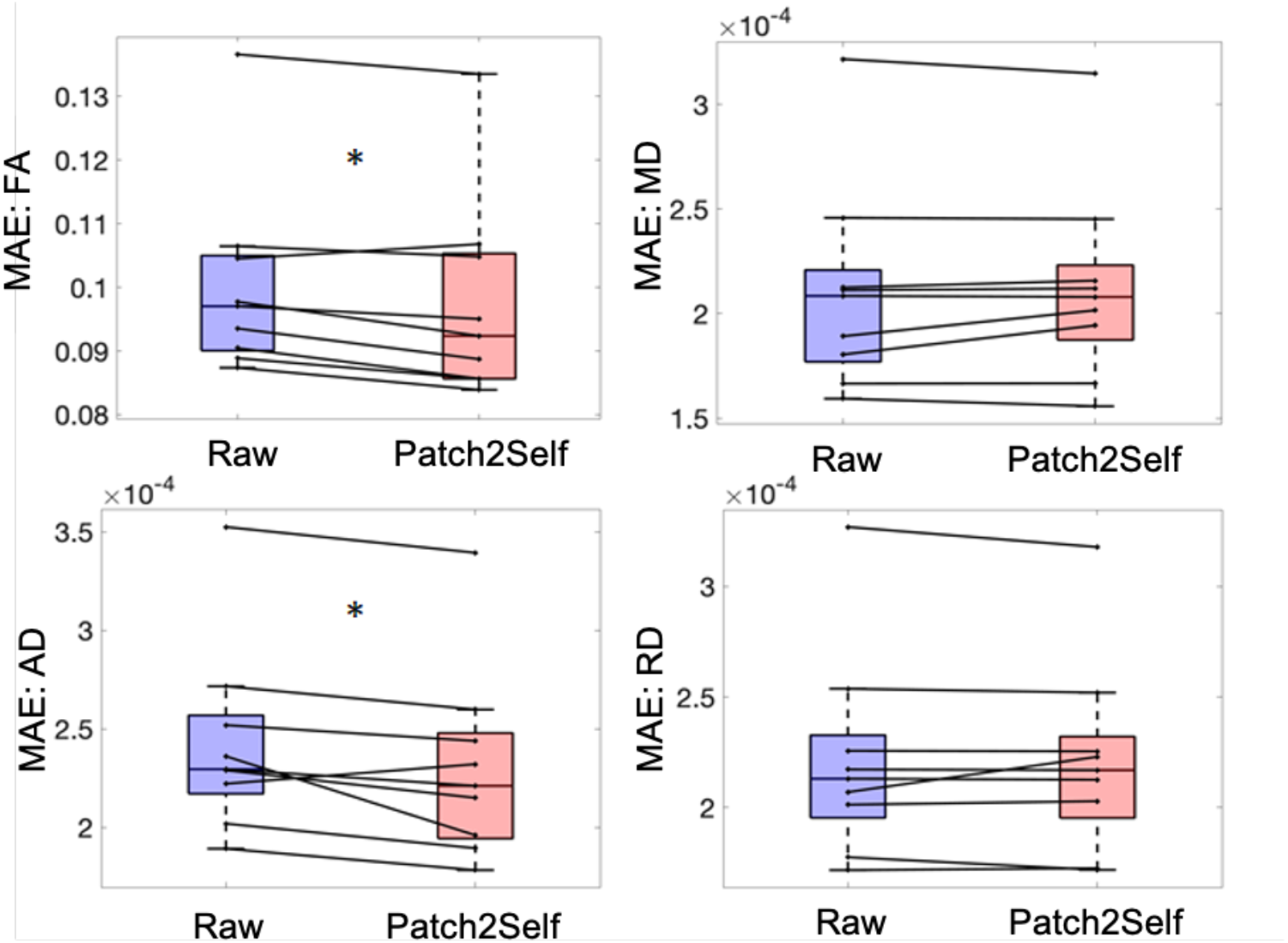
Patch2Self denoising reduces the mean absolute error (MAE) significantly for FA and AD in the scan-rescan dataset. (*p<0.05), indicating improved reproducibility. Denoising does not significantly affect the inter-session variation of MD and RD.

### Averaging Experiments

Cohort B was used to compare Patch2Self on a typical clinically feasible acquisition (∼7 minutes) against averaging several acquisitions over an extended period of time (∼7×5 = 35 minutes). **Figure 7** shows example data from 3 DWIs for 1 to 5 averages (NEX = 1 to 5) along with a single denoised dataset. Qualitatively, and in agreement with theory, averaging improves SNR, with most visually apparent improvement happening after the first averaging operation. Patch2Self denoising shows results qualitatively similar to NEX=4 and NEX=5, with visible noise reduction and no introduction of artificial signal.

**Figure 7:**
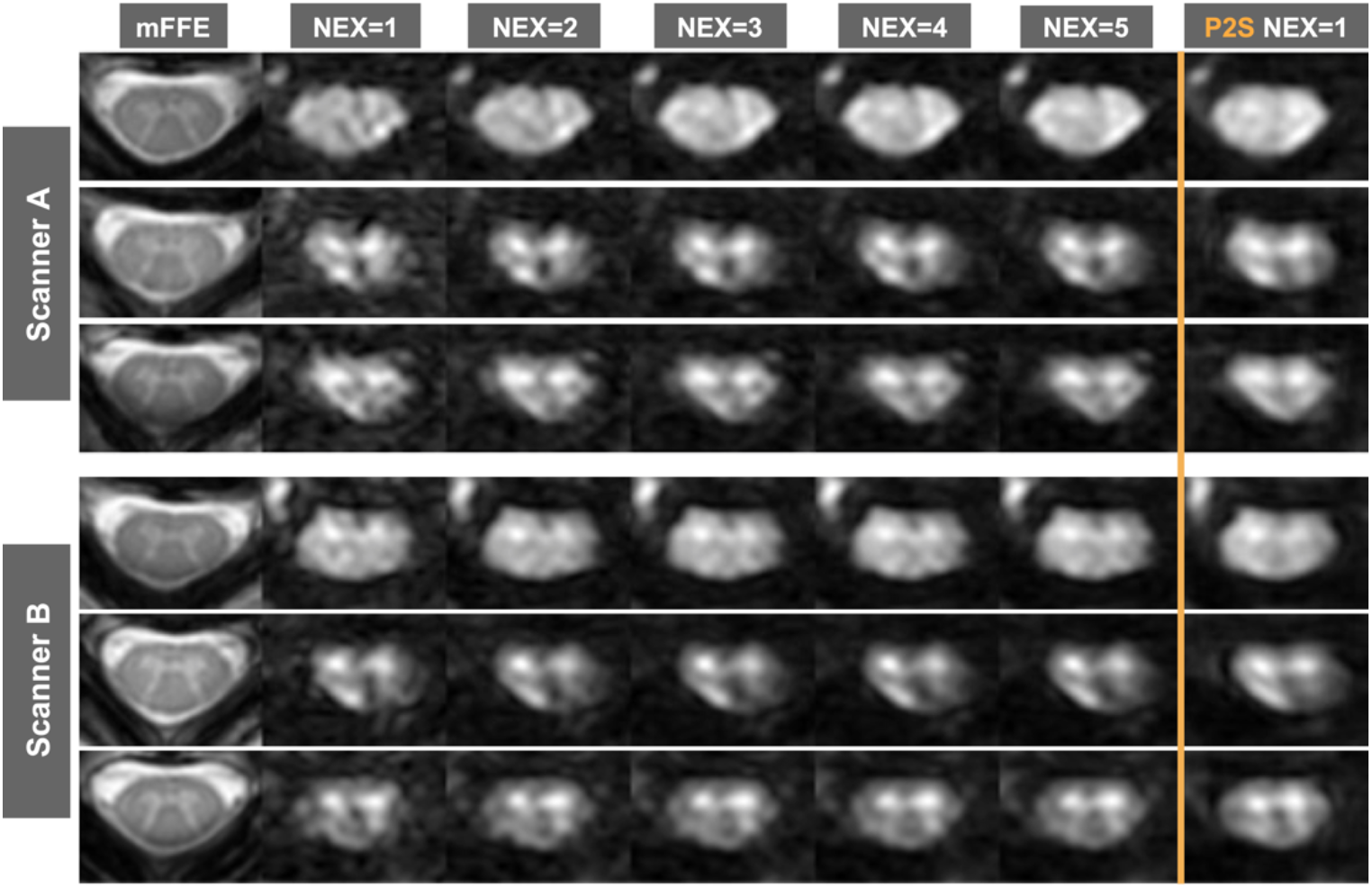
Patch2Self applied to a single dataset (NEX=1, time=7 min) from Cohort B (intra-session repeated acquisitions) qualitatively matches data averaged and acquired in a longer acquisition (up to NEX=5).

SNR is quantified in **Figure 8**. Here, the SNR is shown using raw diffusion data averaged across 1-5 repeats (solid line), where an increase in SNR is confirmed for both scanners. However, a single denoised dataset (dashed line) results in an SNR that is overall higher across a majority of voxels in the cord, for both scanners.

**Figure 8:**
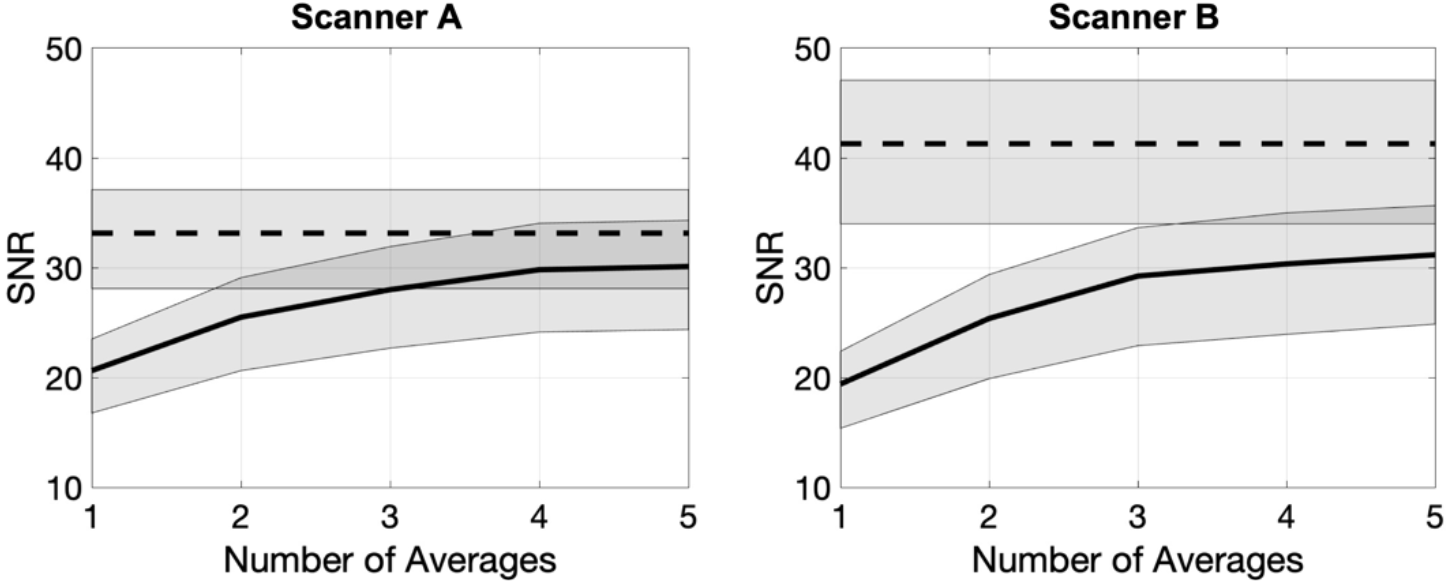
The improvement in SNR with Patch2Self denoising applied to 1 acquisition (dashed lines) is greater than the SNR improvement gained by averaging up to 5 acquisitions (solid lines).

### Patient Experiments

Cohort C consisted of pwRRMS and was used to evaluate lesion conspicuity before and after denoising. **Figure 9** visualizes lesioned spinal cord from two example patients, along with the diffusion signal (from a randomly selected diffusion direction) and DTI-derived measures for both raw and denoised data. While lesions are visible in the raw DWIs, they are more noticeable in the denoised data due to reduced noise variance within WM regions. While not the primary focus of the study, lesions display heterogenous microstructural properties and therefore DTI features. Again, the derived measures show little-to-no changes after denoising, where the greatest benefit is introduced in visualization of the DWIs.

**Figure 9:**
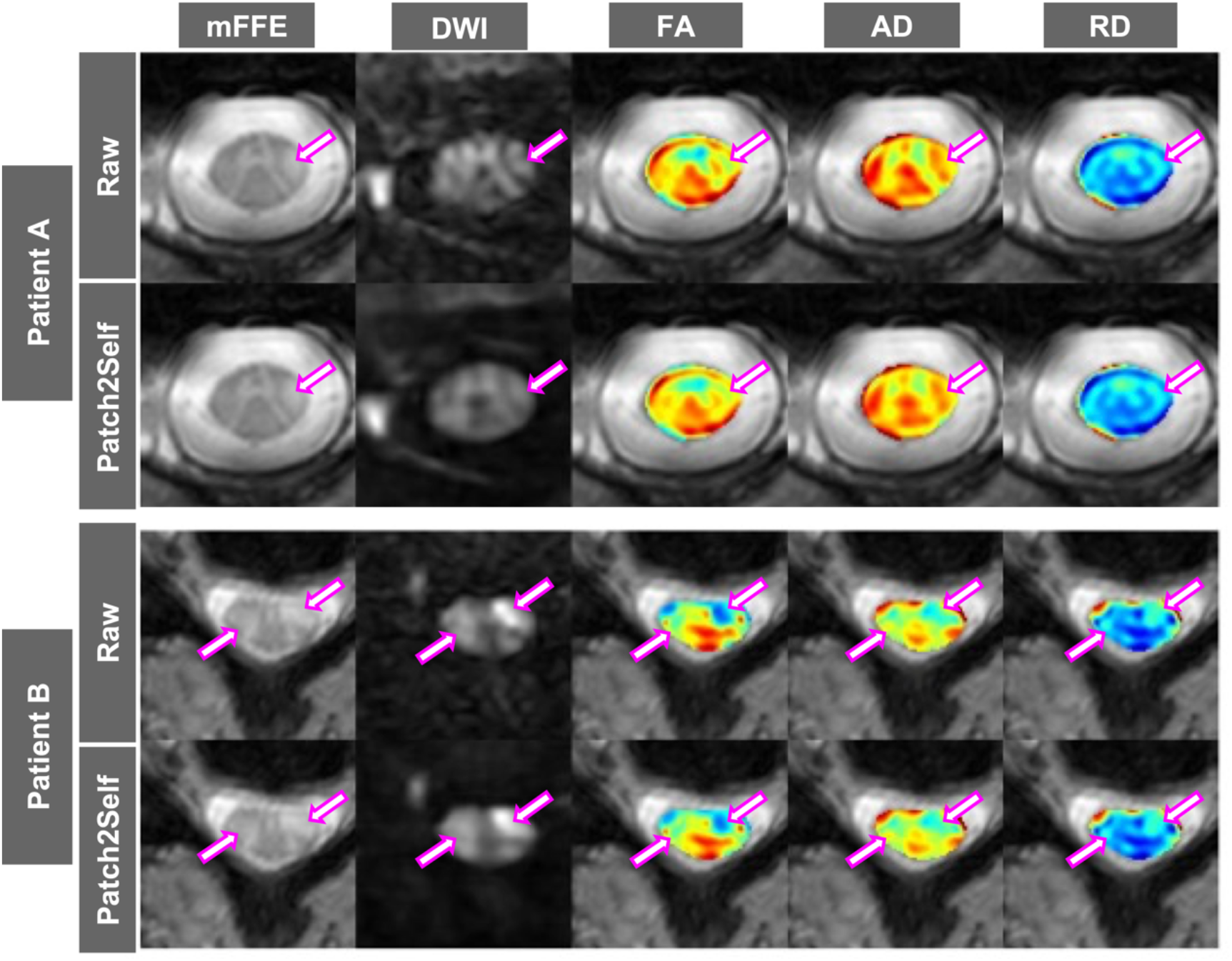
Patch2Self improves conspicuity of lesions in patients with multiple sclerosis. Images from two patients are shown, with both Raw data and data denoised with Patch2Self, where the mFFE, a DWI, and quantitative DTI metrics are visualized for each. Arrows highlight lesions, and qualitative differences in resulting images.

Image quality of pwRRMS data was rated by a trained neuroradiologist, and results quantified in **Figure 10**. Results are shown as a 2D histogram showing ratings before (Raw) and after (Patch2Self) denoising where the same rating would show up on the unity line. Also displayed are the differences in ratings (Patch2Self minus Raw) where a positive value designates improved rating. Patch2Self consistently improved image quality and lesion conspicuity, with most ratings improving by a single rank (or staying the same), and the most common result being an increase in the rating from a 2 to a 3. A two-sided signed rank test confirmed a statistically significant increase (p=9.6E-7) in image rating after denoising.

**Figure 10:**
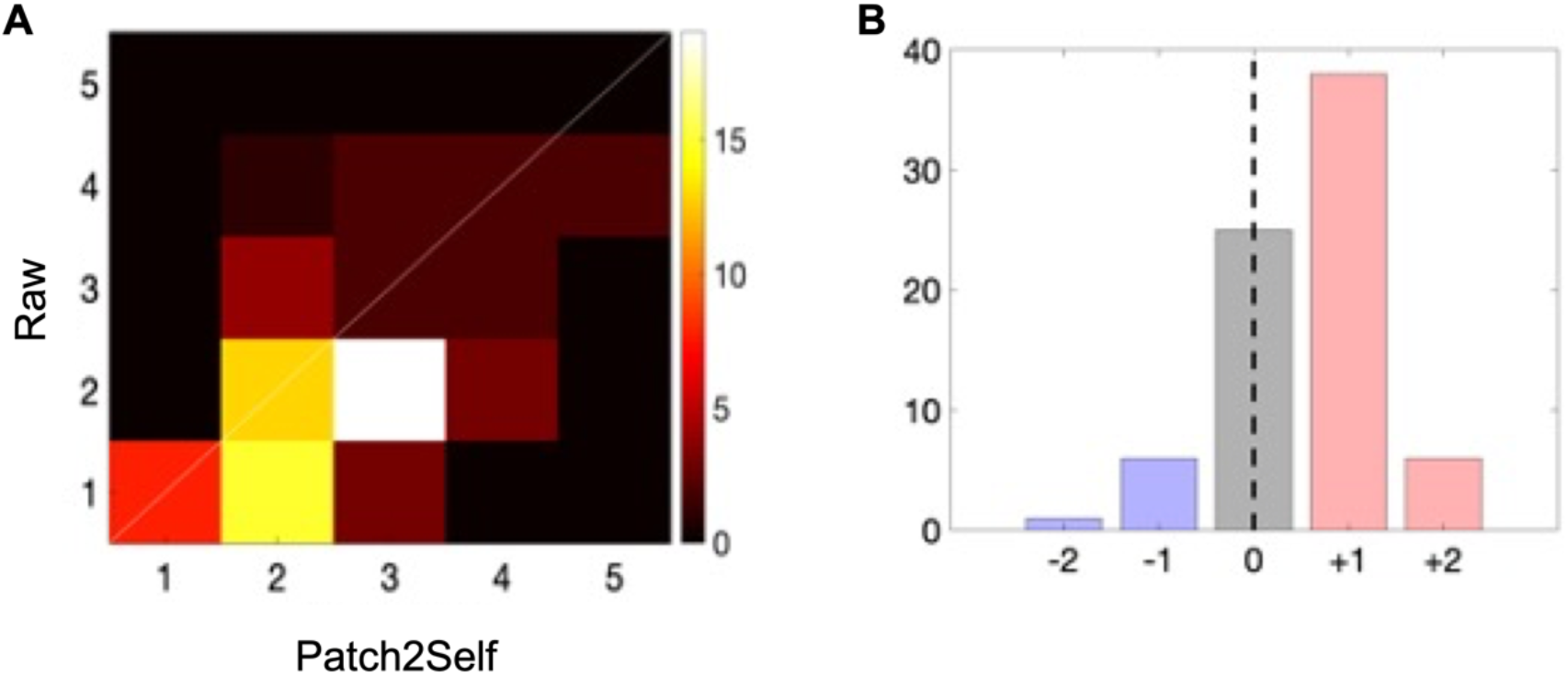
Patch2Self consistently improved image quality and lesion conspicuity in MS patient scans as rated by a trained neuroradiologist. Results are shown as a 2D histogram in (**A**), showing ratings before (Raw) and after (Patch2Self) denoising where the same rating would show up on the unity line. The difference in rating in shown in (**B**) (Patch2Self minus Raw), where a positive value designates improved rating. Most ratings improving by a single value or stayed the same, with the most common result being an increase from a rating of 2 to 3. A two-sided signed rank test confirms a statistically significant increase (p=9.6E-7) in image rating after denoising.

## Discussion

This work demonstrates the advantages of Patch2Self denoising for clinically-feasible in vivo diffusion imaging of the spinal cord. The key findings of this work are that Patch2Self (1) increases intra-cord CNR, (2) improves DTI model fitting, (3) increases scan-rescan reproducibility, (4) increases image SNR, and (5) increases conspicuity of image features and MS lesions in diffusion weighted images. In addition, the Patch2Self algorithm shows no effect of introducing image artifacts or biasing subsequent quantitative diffusion-derived metrics.

Advanced MRI of the cervical spinal cord is challenging due to its small size and mobility, its proximity to tissue interfaces, and its susceptibility to physiological noise from respiratory, cardiac, and pulsatile CSF flow sources. Optimization of acquisition and processing techniques is therefore necessary, as are strategies to increase image quality in post-processing (Rutman et al., 2018). The reliability and sensitivity to pathology of DTI as a quantitative MRI technique make it valuable for various neuroimaging applications.

Spinal cord involvement is a central feature of all MS subtypes and is partly responsible for the accumulation of clinical disability. As a research tool, DTI provides quantitative measures to assess demyelination, edema, fiber integrity and axonal loss in normal-appearing and lesional tissue. DTI metrics have also shown correlations with disability, and with upper and lower limb motor function (Moccia et al., 2019). Reproducible scans with higher SNR will further facilitate such investigations. While MRI is the modality of choice for the management and study of MS, DWI currently has a limited role in the diagnostic work-up (Wattjes et al., 2021). However, given its biological relevance and the breadth of methodological improvements currently ongoing, including in the spinal cord, DTI may soon be a candidate for adoption as an outcome measure instrument in both day-to-day clinical practice as well as clinical trials, especially in progressive MS (Ontaneda et al., 2015). Most importantly, the advantage of this advancement in technology will span over a multitude of spinal cord diseases that diagnosis of which is quite often difficult to reach due to lack of reliable MRI signs. Ischemic spinal cord injury is one of those conditions for which a reliable DTI assessment would permit prompt intervention and change in outcome. The investigation of spinal cord features in health and development, as well as research into acute and chronic injury, degenerative conditions such as neuromyelitis optica spectrum disorder and amyotrophic lateral sclerosis also stand to benefit from improvements in image quality.

Recent harmonization efforts by the research community, including the deployment of optimal DTI acquisition protocols, publication of open-access datasets (Cohen-Adad et al., 2021b), multi-center studies (Cohen-Adad et al., 2021a; Samson et al., 2016) as well as the development of specialized data processing tools (De Leener et al., 2017) will all help promote spinal cord DTI as a valuable research and clinical imaging modality. The present work thus forms part of wider efforts promoting DTI as a promising tool for both research and large-scale clinical applications.

Despite improvements in CNR of the diffusion weighted images, the CNR of DTI-derived metrics did not significantly increase after denoising. This suggests that, with this acquisition protocol, there is already adequate SNR with 15 diffusion images to reliably compute the tensor. Reassuringly, the denoising procedure did not alter the quantified indices, or result in lower CNR, and we expect the reproducibility and CNR of these measures will improve with lower quality acquisitions with lower SNR baseline images.

Overall, the findings in this study support the use of Patch2Self denoising for spinal cord diffusion imaging. While MP-PCA algorithms have become popular in brain imaging (Veraart et al., 2016b), and have proven effective in the cord, even with the use of multiple contrasts (Grussu et al., 2020), data redundancy is required. In this case, the datasets are limited to low-direction, single b-value images, and application of MP-PCA to these data shows little-to-no difference in diffusion images with our 15-direction datasets. Future work should investigate different denoising techniques on datasets tailored for advanced multi-compartment modeling, which are expected to have redundant data, and their effect on contrast and derived measures of tissue microstructure.

### Limitations

The goal of this study was to provide a demonstration of the potential benefits of applying a denoising technique to diffusion MRI of the spinal cord. Some limitations include the relatively small sample sizes of healthy volunteers and pwMS, and the use of a single diffusion acquisition protocol and model (DTI). Future studies would benefit from exploring the effects of the Patch2Self denoising method for a range of acquisition parameters and different model approaches (e.g., models for multi-shell diffusion data). In addition, our cohort of pwMS was formed by relatively young subjects with minimal degree of disability. These factors contributed to minimizing motion artifacts which may be more commonly seen when imaging pwMS with more advanced disease or pain. While only one MRI scanner vendor (Philips) was used in this study, data sets from two different scanners with differing gradient performance specifications were analyzed. With respect to the rating of patient diffusion image quality by a neuroradiologist, we used individual DWIs rather than the mean DWI or mean apparent diffusion coefficient image, which differs somewhat from standard clinical workflow. When individual directions are viewed, a lesion is not always visible. However, the goal of this experiment was to provide an initial evaluation of perceived image quality with a larger sample size than could be achieved with a single mean image per patient. Additionally, dMRI preprocessing in the spinal cord has not been fully optimized (Snoussi et al., 2021), and future work should investigate the effects of denoising algorithms in combination with motion and distortion correction methods.

## Conclusions

Here, we have shown that the application of Patch2Self denoising in clinical-quality spinal cord diffusion data improves intra-cord contrast, signal fitting, SNR, and lesion conspicuity. This denoising approach is freely available in the DIPY software package and can be implemented on any in vivo dMRI spinal cord acquisitions. This algorithm and approach hold promise for facilitating reliable diffusion measures in the spinal cord to investigate biological and pathological processes.

## Data Availability

The datasets supporting the conclusions of this article will be made available by the authors upon request.

## Acknowledgements

The authors thank the Vanderbilt University Institute of Imaging Science (VUIIS) Center for Human Imaging, the VUIIS technologists, and all study participants. Research reported in this publication was supported in part by funding from the National Institutes of Health under award numbers 5R01NS109114 (S.A.S.), R01NS117816 (S.A.S.), R01EY023240 (S.A.S.), R01EB017230 (B.A.L.), R01EB027585 (S.F. and E.G.), KL2TR002245 (K.P.O.), and K01EB030039 (K.P.O.), by the National Multiple Sclerosis Society award number RG-1501-02840 (S.A.S.), the Conrad Hilton Foundation (S.A.S.) and in part by the National Center for Research Resources grant UL1RR024975-0. Francesca Bagnato receives research support from Biogen Idec, the National Multiple Sclerosis Society (RG-1901-33190), the National Institutes of Health (R21NS116434-01A1) and the Veterans Health Administration (1 I01 1 I01 CX002160-01).

